# A Swiss *DSG2* Founder Variant Promotes Left Ventricular Thrombus Formation Causing Cardioembolic Stroke in Autosomal Recessive Arrhythmogenic Cardiomyopathy

**DOI:** 10.64898/2026.08.17.26359854

**Authors:** Sandra A. Hemkemeyer, Stefania Quintiliani, André Schaller, Raouf Madkhour, Elena Elchinova, Jennifer Schröder-Schwarz, Pauline Hanns, Christiane Zweier, Katja E. Odening, Camilla Schinner, Marina Rieder

## Abstract

**Aims:** Arrhythmogenic cardiomyopathy (ACM) is a genetic disease defined by arrhythmias and myocardial fibrosis with impaired cardiac function and increased risk of sudden cardiac death. Pathogenic variants are mostly identified in desmosomal genes such as desmoglein-2 (*DSG2*). We identified a novel disease phenotype in patients homozygous for the *DSG2* variant c.523+2T>C (splice site of exon 5/intron 5), characterized by cardioembolic events in addition to classical ACM features. Here, we evaluate this new thromboembolic phenotype by comparing the clinical data to specific murine disease models.

**Methods and Results:** We describe three unrelated patients presenting with an embolic event and/or left ventricular thrombus. Clinical evaluation revealed a shared right ventricular ACM phenotype characterized by arrhythmias, impaired function, and fibrotic remodeling. In addition, patients exhibited localized fibrotic changes of the left ventricular apex with formation of an aneurysm and predisposition to thrombus formation. Genetic analysis identified the *DSG2* variant c.523+2T>C as a Swiss founder variant. To elucidate the variant’s functional impact, a mouse model deficient for Dsg2 exon 5 (Dsg2Δex5) was established and compared to a model carrying the adhesion-deficient Dsg2-W2A variant. Echocardiography, ECG, and histology in Dsg2Δex5 mice revealed similar disease patterns to patients and a loss of DSG2 expression. Importantly, these animals exhibited left apical fibrosis with aneurysm formation and left ventricular thrombus formation. In contrast, the Dsg2-W2A model presented with a biventricular ACM-phenotype, but without left ventricular thrombi.

**Conclusions:** We identified a novel ACM phenotype in patients homozygous for the *DSG2* founder variant c.523+2T>C characterized by left ventricular apical fibrosis. Dsg2Δex5 mice recapitulate the patients’ phenotype suggesting a causative link between left ventricular aneurysm due to DSG2 deficiency and thrombus formation with subsequent embolism. This highlights a novel pathological feature of ACM and the need for variant- and phenotype-specific therapy.

**Clinical Perspective:** *What Is New?:*

- Identification of a Swiss *DSG2* founder variant associated with left ventricular apical fibrosis, thrombus formation and increased risk for cardioembolic ischemic strokes
- First evidence of variant- and phenotype-specific therapy in *DSG2*-associated arrhythmogenic cardiomyopathy

*What Are the Clinical Implications?:*

- Homozygous carriers of specific *DSG2* variants may require prophylactic anticoagulation once left ventricular apical fibrosis is detected.
- Precision medicine approaches in arrhythmogenic cardiomyopathy should consider variant-specific phenotypes.

## Introduction

Arrhythmogenic right ventricular cardiomyopathy (ARVC) is a heritable myocardial disorder characterized by progressive replacement of myocardium with fibrofatty tissue, leading to ventricular arrhythmias and an increased risk of sudden cardiac death (1, 2). Initially described as a disease predominantly affecting the right ventricle, advances in clinical evaluation and imaging have revealed a broader phenotypic spectrum, including left-dominant and biventricular forms. This evolving understanding has prompted a shift in disease classification from ARVC to the more inclusive term “arrhythmogenic cardiomyopathy” (ACM) (3). Beyond arrhythmias, heart failure resulting from progressive myocardial dysfunction has also emerged as a significant clinical manifestation of ACM (4).

Pathogenic variants in desmosomal genes — such as plakophilin-2 (*PKP2*), desmoplakin (*DSP*), plakoglobin (*JUP*), desmoglein-2 (*DSG2*), and desmocollin-2 (*DSC2*) — are the primary genetic drivers of ACM (5). Pathogenic variants in *PKP2* are the most prevalent, accounting for the majority of genetically characterized cases (6–8). *PKP2* encodes a plakophilin protein crucial for anchoring intermediate filaments to the desmosomal plaque, thereby preserving cardiomyocyte structural integrity (9). In contrast, variants in *DSG2* and *DSC2* — both desmosomal transmembrane proteins essential for cell-cell adhesion and mechanical cohesion during cardiac contraction - are less common, accounting for approximately 5 – 15 % of cases depending on the geographic population (6–8, 10). While *PKP2*-associated ACM has been extensively studied, comprehensive cohort data on *DSG2*- and *DSC2*-related disease have only recently become available (11). Like patients with the “classical” *PKP2*- associated ARVC, patients with variants in *DSG2* present with predominantly right ventricular involvement, while biventricular disease is mainly described for clinically advanced forms. Recent data revealed that approximately one third of the individuals with *DSG2*- or *DSC2*- associated ACM carry multiple variants, either in a homozygous/compound heterozygous state or as digenic heterozygotes (11). Consequently, in contrast to the predominantly autosomal dominant inheritance pattern observed in *PKP2*-associated disease, *DSG2*-associated ACM may, in part, follow an autosomal recessive mode of inheritance (11). Accordingly, previous reports have described the presence of founder variants, suggesting a geographically or ethnically driven accumulation of specific variant (10, 12). Patients with autosomal recessive ACM due to bi-allelic variants exhibit higher penetrance and earlier disease onset, as well as an increased risk of adverse arrhythmic events and heart failure outcomes compared to individuals with heterozygous variant (11).

While the genetic and main clinical features of *DSG2*-associated ACM are becoming increasingly clear, certain complications remain rare and less characterized. Among these, thrombus formation is an important clinical phenomenon. In ACM, thrombi predominantly affect the right ventricle and typically occur in the context of severe ventricular dysfunction or additional cardiac comorbidities. Notably, left ventricular thrombi are exceptionally rare in classical ACM, particularly when left ventricular (LV) systolic function is preserved or only mildly impaired (13, 14).

In this study, we identified a *DSG2* splice-site variant (c.523+2T>C) as a Swiss founder variant in a homozygous state in three unrelated patients who developed left ventricular thrombi and/or strokes as a consequence of ACM.

To further explore the underlying mechanisms of this new ACM phenotype, we generated a mouse model with inducible deletion in the same genetic region as the patients’ variant (Dsg2Δex5) and compared its phenotype to the Dsg2-W2A knock-in model, a previously described distinct *Dsg2* loss-of-function mutation leading to defective desmosomal adhesion (15). *In vivo* echocardiography and electrocardiogram (ECG) assessments were performed, followed by detailed histological analysis. The experimental findings in the mouse model mimic the clinical disease features of patients homozygous for the Swiss founder *DSG2* variant including the presence of left ventricular fibrosis and thrombus formation providing important mechanistic insights into focal myocardial injury, fibrosis, and thrombogenesis linked to a *DSG2* homozygous genotype.

## Materials and Methods

### Clinical Data

We describe the cases of three unrelated patients who were evaluated and treated at the Department of Cardiology, Inselspital, Bern University Hospital (Switzerland), due to ACM. All cases underwent comprehensive diagnostic work-up, including ECG, transthoracic echocardiography (TTE), cardiac magnetic resonance imaging (MRI), Holter monitoring, and genetic testing by panel analysis. The clinical diagnosis of ACM was made in accordance with the 2010 revised Task Force Criteria (16). The patients provided written informed consent for the publication of clinical data, and also for further analysis of their sequencing data to investigate a potential founder effect (Cantonal Ethics Board Bern, 2021-01396).

### Molecular Genetic Testing

Blood samples were taken from patients, and genomic DNA was extracted from blood leukocytes according to manufacturer instructions (for patient 1: Maxwell RSC Blood DNA Kit, Promega, Madison, USA and for patients 2 and 3: Prepito DNA Blood250-Kit, PerkinElmer, Massachusetts, USA). Genetic testing for pathogenic variants in patient 1 and 2 was performed using whole exome sequencing (Twist Comprehensive Exome, Twist Bioscience, San Francisco, USA) on a NovaSeq 6000 instrument (Illumina, San Diego, USA). Patient 3 was sequenced using TruSight One enrichment kit (Illumina, San Diego, USA) on a MiSeq instrument (Illumina, San Diego, USA).

### Haplotype analysis

To investigate whether the patients have an identical homozygous haplotype (ROH) surrounding *DSG2*, which would suggest that this *DSG2* variant is a founder variant, sequencing data were analysed using the AutoMap homozygosity mapping tool (17).

### Animal Experiments

All animal experiments were carried out according to the protocol approved by the Cantonal Veterinary Office of Basel-Stadt (License number 3070) and complied with the ARRIVE guidelines. All mice were housed under specific pathogen-free conditions with standard chow and bedding with 12 hours day/night cycle according to institutional guidelines. Both male and female animals were included without sex-based selection.

### Animal Models

To generate Dsg2Δex5 mice, animals of the B6.129*Dsg2*tm1Mdcb strain (18, 19) (kindly provided by the Max-Delbrück-Centrum für Molekulare Medizin in der Helmholtz-Gemeinschaft, Arnd Heuser) which harbor *Dsg2* exon 5 flanked by *loxP* sites (floxed), were crossed with B6.Cg-Tg(CAG-cre/Esr1*)5Amc/J animals (004682, The Jackson Laboratory, Bar Harbor, USA), which harbor the Cre transgene under a tamoxifen-inducible CAG-promotor. For CAG-Cre induction, tamoxifen (Cat# T5648, Sigma-Aldrich) was dissolved at 20 mg/ml in corn oil (Cat# C8267, Sigma-Aldrich) and animals were injected with tamoxifen for 5 consecutive days at a concentration of 75 mg/kg body weight. ‘CRISPR/Cas9-based gene editing and detailed phenotyping of the Dsg2-W2A knock-in mouse model as previously described (15).

### Genotyping

For genotyping of both mouse lines, DNA was extracted from tissue biopsies in 25 mM NaOH/0.2 mM EDTA at 98 °C for 1 hour and neutralized by adding an equal volume of 40 mM Tris HCl pH 5.5. PCR was performed using GoTaq Flexi (M7845, Promega, Madison, WI, USA) according to manufacturer’s instructions with primers at final concentration of 1 µM. For the Dsg2Δex5 line, the primers 5’-CCAGAGGAAACAACCTGGAA-3’ (forward) and 5’- GCACAGGACTCAGGATTGGT-3’ (reverse), which span part of the floxed region of *Dsg2*, and 5’-GCTAACCATGTTCATGCCTTC-3’ (forward) and 5’-AGGCAAATTTTGGTGTACGG-3’ (reverse) for detection the CAG-Cre transgene were applied. For the Dsg2-W2A line, the following primers were used: 5’-GAATGTCTCCCCAAAGCTTTGGGTATG-3’ (forward) and 5’- CTGCTACCTTGGCAATCGGGTTC-3’ (reverse), which span the mutated region. For Dsg2-W2A, the PCR product was restricted with 66.7 U/ml *Alu*I (R0137, New England Biolabs, Ipswich, MA, USA) in CutSmart buffer (New England Biolabs) at 37 °C for 1h. Electrophoresis of all PCR products was performed according to standard procedures using agarose/TAE gels and Midori Green Advance (Cat# MG04, Nippon Genetics, Düren, Germany) for fluorescence DNA visualization.

### Echocardiography and ECG

Transthoracic echocardiography was performed using a Vevo 2100 high-resolution ultrasound system (VisualSonics, Toronto, Canada) equipped with an MS-550 linear-array transducer operating at a central frequency of 40 MHz. Mice were anesthetized with 3.0% (v/v) isoflurane in compressed air and positioned in a supine position on a pre-warmed imaging platform, and anesthesia was maintained with 1.5% (v/v) isoflurane delivered via a nose cone. Body temperature was continuously monitored using a rectal thermocouple probe and maintained at 37 °C. Chest hair was removed using a commercially available depilatory cream (Nair) to ensure optimal acoustic coupling. Cardiac geometry and function were assessed using B-mode imaging in parasternal long-axis (PLAX) and short-axis (SAX) views at the mid-papillary muscle level. For tricuspid annular plane systolic excursion (TAPSE) apical 4-chamber view was acquired.

Right ventricular (RV) output function was evaluated in B-mode SAX view by measuring RV fractional area change (RV FAC). RV endocardial borders were traced at end-diastole and end-systole. In addition, TAPSE was measured, and the right ventricular outflow tract (RVOT) diameter was determined in the PLAX view. Left ventricular (LV) function was assessed in the parasternal short-axis view at the mid-papillary muscle level. LV anterior and posterior wall thickness as well as internal dimensions were measured using 2D-guided M-mode imaging, and LV volumes and LV ejection fraction (LVEF) were calculated using the Teichholz formula. In parallel, LV systolic function was independently evaluated from B-mode images by tracing the endocardial area at end-diastole and end-systole, and LVEF was determined using area-based calculation. All echocardiographic measurements were performed by a single observer to minimize inter-operator variability, and data were analysed using Vevo 2100 software (version 1.6.0, VisualSonics).

For ECG recordings, mice were kept anesthetized after echocardiography and directly transferred onto a pre-warmed heating pad in supine position and attached to the PowerLab Data Acquisition System (ML870 Powerlab 8/30, ADInstruments, Sydney, Australia) via insertion of subcutaneous needle probes into the right upper, and both lower limbs for acquisition of lead II. ECG was recorded for 5 min. Recording and analysis of ECG data was performed using the LabChart Pro 8 software (ADInstruments) equipped with the ECG Analysis Module. All mouse data were analysed by an investigator blinded to the study groups. From three regions, 100 consecutive QRS complexes were averaged and quantified, applying the following recognition settings: Typical QRS width 10 ms, R waves at least 60 ms apart, pre-P baseline 10 ms, maximum PR 50 ms, maximum RT 60 ms. For each animal, the values of the parameters were averaged across the three regions and plotted. After final measurements, mice were euthanized via cervical dislocation under anaesthesia, and the hearts were collected as described below.

### Murine organ collection

For heart dissection, mice were euthanized via cervical dislocation under isoflurane anaesthesia according to guidelines of the Cantonal Veterinary Offices and Universities. Hearts were removed by lateral thoracotomy and directly immersed in ice-cold HBSS supplemented with 20 mmol/l 2,3-Butanedione monoxime (BDM, B0753, Sigma-Aldrich). Hearts were fixed in 4 % paraformaldehyde/PBS at 4°C overnight and evaluated macroscopically including acquisition of images from the anterior facies with a Zeiss Stemi 508 binocular equipped with an Axiocam 208 Color camera. Subsequently, hearts were processed for formalin-fixed paraffin embedding (FFPE) employing a Leica TP 1020 embedding device (50% Isopropanol 90 min, 70% Isopropanol 30 min, 70% Isopropanol 90min, 90% Isopropanol 60 min, twice 90 % Isopropanol for 120 min each, trice absolute Isopropanol for 210 min each, twice paraffin 5h each; all steps at room temperature, except paraffin at 63°C). Murine brain was harvested from an animal that pre-maturely died during the experiment, morphology was documented by Zeiss Stemi binocular as described above and then embedded in embedded in O.C.T. compound (Cat# SA62550-01, Tissue-Tek). Brain cross-section was obtained by cutting the tissue block using a ThermoFisher Cryostar NX50.

### Histological staining

FFPE samples were sectioned using a microtome (Microm HM 430, Epredia, Thermo Fisher Scientific) at a thickness of 4 μm, transferred to SuperFrost Plus slides (Thermo Fisher Scientific) and dried at 37°C overnight. Hematoxylin / Eosin (H.E.) staining was performed using a Shandon Varistain 24-4 Slide Stainer (ThermoFisher Scientific) according to standard procedures. In brief, FFPE sections were deparaffinized and stained with Mayer’s hemaluan solution (Cat# 1.09249, Sigma-Aldrich) for 3 min, washed, and stained with 0.5 % (w/v) eosin solution (Erythrosin B, Cat# 198269-25G, Sigma Aldrich) for 3 min. After washing in water, sections were dehydrated in an ascending ethanol row and xylene substitute HistoClear II XEM 200 (Cat# ND-HS-200, Vogel), sections were mounted using Roti-Histokit II (Cat# T160.2, Roth).

Sirius red staining was performed according to standard procedures. In brief, FFPE sections were deparaffinized, washed in distilled water and stained with Coelestine blue iron-alum solution (Cat# 15156.00500, Morphisto, Offenbach aM, Germany) for 7 min, followed by Mayeŕs hemalum solution (Cat# 1.09249.0500, Sigma-Aldrich) for 7 min, differentiated in running tap water and stained with Picro-Sirius Red solution (Cat# 13422.00500, Morphisto) for 30 min. Sections were dehydrated in an increasing ethanol series and cleared in HistoClear II XEM 200 and mounted in Roti-Histokit II. Images of histological sections were acquired with a 20x objective mounted on a AxioScan Z1 slide scanner (Zeiss, Jena, Germany). The area of collagen was analysed using QuPath software 0.5.1. Areas of interest (i.e. apex, interventricular septum, right and left ventricles) were annotated in a blinded manner, color channels were separated by deconvolution and total and fibrotic tissue area was determined by applying the “fibrosis analysis” script (15).

### Immunostaining

Immunostaining was performed according to standard procedures. Briefly, FFPE sections were deparaffinized and temperature-mediated antigen retrieval was performed in 10 mM Tris, 1 mM EDTA pH 9, 0.05 % Tween at 191 °C for 10 min. Tissue was permeabilized in 0.1 % Triton X-100 in PBS for 5 min and blocked with 3 % bovine serum albumin (BSA)/0.1 % normal goat serum in PBS for 1 hour.

The following primary antibodies were incubated in 1 % BSA/PBS at 4 °C overnight: anti-Fibrin (Cat# ZMS1211, stock 0.3 mg/mL, 1:100, Sigma), anti-desmoglein-2 (Cat# 610121, 1:100, Progen), anti-N-Cadherin (Cat# 610921, 1:200, BD BioScience). Respective isotype controls were applied to neighbouring sections (mouse IgG1, Cat# 02-6100, stock 1 mg/ml, 1:333, Invitrogen; rabbit negative control, Cat#X0903, DAKO, 1:1000). Slides were washed and incubated with respective secondary goat anti-mouse or anti-rabbit antibodies coupled to Alexa Fluor 488 or Alexa 568 (Cat# A11008, Cat# A11004, Cat# A11001, 1:300 all Invitrogen) diluted in 1% BSA/PBS and incubated for room temperature for 1 hour. To visualize the cell membrane, wheat germ agglutinin (WGA)-Alexa Fluor 647 (Cat# W32466, Invitrogen) was incubated together with the secondary antibody. DAPI (Cat# D9542, Sigma-Aldrich) was added with a final concentration of 1 µg/ml for 10 minutes to counterstain nuclei. After washing in PBS, sections were mounted with Fluoromount Aqueous Mounting Medium (Cat# F4680, Sigma-Aldrich). For wide field image acquisition, a 40x objective mounted on a AxioScan Z1 slide scanner (Zeiss, Jena, Germany) was employed and for confocal image acquisition a Leica SP8 microscope equipped with HCX PL APO CS 10x/0.40 DRY or HC PL APO CS2 63x/1.30 GLYC objectives and Leica LAS-X software was used.

### Statistics and image compilation

Figures were compiled with Adobe Illustrator CC 2025. Statistical computations were performed with Prism 9 and 10 (GraphPad Software, Boston, USA). For comparison of two or multiple groups, distribution of data was analysed by a Shapiro-Wilk normality test revealing a non-normal distribution for all data sets. Group variances were analysed by non-parametric Kruskal-Wallis with Dunn’s post-hoc test. Statistical significance was assumed at p < 0.05, respective p-values are indicated in the figures. Box plots indicate the median and 25^th^ to 75^th^ percentiles with whiskers from minimum to maximum. Each animal was considered as a biological replicate.

## Results

### Clinical Data

#### Patient 1

A woman in her 30’s and with unremarkable medical history presented with sudden left-sided hemianopsia and occipital headache. She did not engage in high-intensity sports, and her family history showed no significant cardiovascular disease. A cerebral MRI revealed an ischemic stroke in the right posterior cerebral artery (PCA) territory with complete demarcation. The patient was admitted to the stroke unit and initially treated with aspirin, as the thrombolysis window had already passed.

As part of the basic diagnostic workup, a resting ECG was performed and showed global low QRS voltage with QRS amplitudes <5 mV in all limb leads and <10 mV in all precordial leads in conjunction with anterior T-wave inversions (**Figure 1A**). 24-hour Holter ECG monitoring revealed 2900 polymorphic premature ventricular contractions (PVCs) and one episode of non-sustained ventricular tachycardia (3 consecutive beats, 117/min), while no atrial fibrillation was detected.

**Figure 1.**
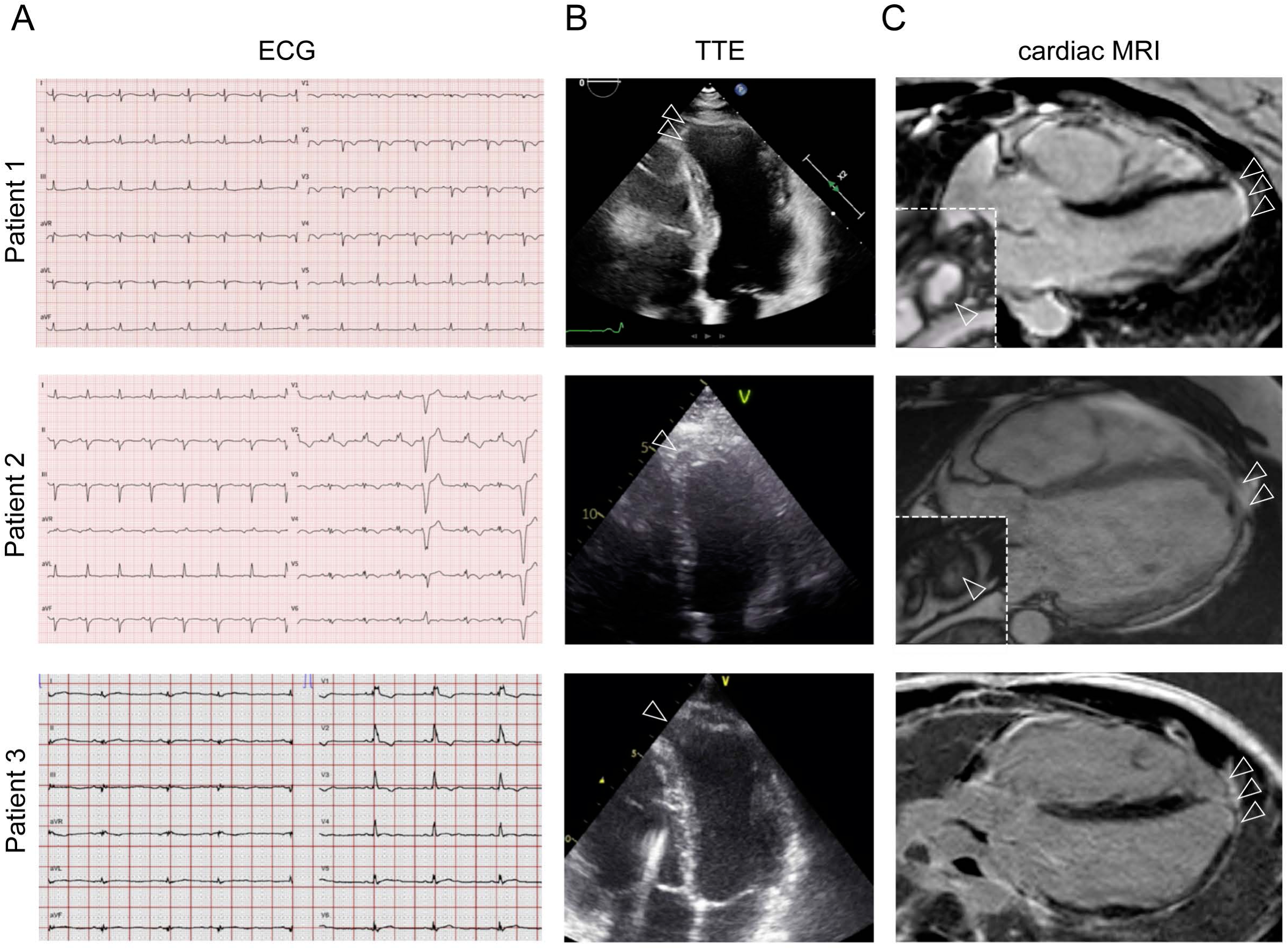
Clinical findings in patients. (A) Resting ECG at initial presentation recorded at 25mm/sec and 10mm/mV. (B) Transthoracic echocardiographic (TTE) images of the left ventricle showing an apical aneurysm (arrowheads) for patient 1 – 3. (C) Cardiac MRI demonstrating transmural late gadolinium enhancement (LGE) in the left ventricular apex (arrowheads) with visualization of a small thrombus in the LV apex (insert, arrowheads) for patient 1 (5 x 5mm) and patient 2 (4 x 8mm).

To further investigate the etiology of the ischemic stroke and of the ECG findings, both TTE and transesophageal (TOE) echocardiograms were performed. These revealed impaired RV function, with a TAPSE of 16 mm and a TV annulus DTI S’ of 7.5 cm/s, alongside RV dilation (PLAX RVOT 39 mm, 21.6 mm/m²). Additional findings included apical hypertrabeculation and hypokinesia of the RV free wall. A localized aneurysm with dyskinesia at the left ventricular (LV) apex was also observed, although the LVEF was globally normal (LVEF 60 %) (**Figure 1B; Supp. Video 1**). No evidence of an intracardiac thrombus or a patent foramen ovale was found on echocardiography.

Cardiac MRI confirmed normal LV size and function (LV end-diastolic volume, LV EDV 125 ml, LVEF 59 %). Consistent with the echo findings, it further revealed transmural late gadolinium enhancement (LGE) with akinesia of the mid-ventricular lateral wall and apex, accompanied by a small apical aneurysm containing a thrombus (5 x 5 mm) (**Figure 1C**). The RV was dilated (RV EDV 101 ml/m²) with impaired systolic function (RVEF 35 %), along with fibrosis and consecutive dyskinesia in the subtricuspid region, and hypokinesia extending from the mid-ventricular to the apical free wall.

In conclusion, the diagnosis of ACM was established according to the 2010 task force criteria (16) and the small LV thrombus due to the LV apical scar was considered the cause of the stroke. Oral anticoagulation with rivaroxaban was initiated. To confirm that the LV findings were due to ACM rather than an ischemic cause, coronary CT angiography was performed five days after the initial cardiac MRI, which revealed normal coronary arteries and complete resolution of the thrombus. A blood coagulation screening was conducted and revealed no abnormalities. Genetic testing by panel analysis revealed that the patient was homozygous for the variant c.523+2T>C, p.? in the *DSG2* gene. The reported splice-site variant was classified as pathogenic according to the American College of Medical Genetics criteria (20). It affects the highly conserved splice donor of intron 5 and is predicted to result in aberrant splicing by exon 5 skipping, thus leading to a frameshift and subsequent degradation of the mutant transcript via nonsense-mediated mRNA decay. Sequencing data did not indicate a deletion at this locus, making hemizygosity of the variant highly unlikely. Notably, the parents were not knowingly related for more than three generations, and there was no reported history of cardiac disease or sudden cardiac death in the family.

#### Patient 2

A man in his early 40’s with no relevant previous medical history called an ambulance due to presyncope, dizziness, and chest pain. The rhythm monitoring conducted by the paramedics revealed a broad complex tachycardia with ventricular-atrial dissociation at a heart rate of 250 beats per minute, which spontaneously resolved.

The subsequent 12-lead ECG, after normalization of the heart rate, showed a right bundle branch block, anterior T-wave inversion, low-voltage in leads V3-V6, and polymorphic PVCs (**Figure 1A**).

TTE revealed LV apical akinesia, RV dilatation with impaired function (FAC 29 %), and regional RV wall motion abnormalities (**Figure 1B, Supp. Video 2A and 2B).** Holter ECG monitoring demonstrated a PVC burden of 7,250 per 24 hours and 180 episodes of non-sustained ventricular tachycardia (maximum of 3 consecutive beats).

Cardiac MRI confirmed the findings from the TTE, revealing RV dilatation (RV EDV 120 ml/m²), impaired RV function (RVEF 28 %), and regional akinesia or dyskinesia of the fibrotic lateral RV wall. Additionally, LV dilatation (LV EDV 104 ml/m²), reduced LV function (LVEF 39 %), and an apical aneurysm with a small thrombus (4 x 8 mm) were observed (**Figure 1C**).

The diagnosis of ACM was established according to the 2010 task force criteria (16) and oral anticoagulation with apixaban due to LV-thrombus, beta-blockers, and heart failure therapy was initiated and the patient received an implantable cardioverter-defibrillator (ICD) for secondary prevention.

Panel analysis revealed the homozygous variant c.523+2T>C, p.? in *DSG2*, even though the parents were not knowingly related to each other. Other than an unexplained death of an elderly family member, the family history was unremarkable. A blood coagulation screening revealed no abnormalities.

A few years after the initial presentation, the patient experienced an electrical storm, consisting of 17 episodes of sustained ventricular tachycardia, successfully treated with ATP, and 3 episodes that required appropriate ICD shocks due to failed ATP. Following a targeted epicardial VT ablation in the inferior basal region of the RV epicardium, the patient has not had any further episodes to date.

#### Patient 3

During a routine medical evaluation, an adolescent male with no known relevant medical history was incidentally found to have conduction and repolarization abnormalities on a 12-lead ECG, including right bundle branch block, anterior T-wave inversions, and left anterior fascicular block (**Figure 1A**). The parents were not knowingly related to each other, and there was no family history of cardiovascular disease. The patient reported normal exercise tolerance and had never experienced syncope.

TTE revealed RV dilation with impaired systolic function (TDI 8.1 cm/s, TAPSE 19 mm, FAC 29 %). Additionally, mild global LV dysfunction was observed (LVEF 50 %) with diffuse hypokinesia and a small, localized dyskinetic area at the left ventricular apex (**Figure 1B**). Cardiac MRI confirmed these findings and demonstrated mildly reduced LV systolic function (LVEF 52 %) with transmural LGE localized to the LV apex (**Figure 1C**). The right ventricle was severely dilated (RV end-diastolic volume index [RV EDVI] 223 mL/m²) with significantly impaired function (RVEF 27 %), marked wall thinning, and regional wall motion abnormalities affecting the RV free and inferior walls. LGE as a sign of fibrosis was also present in the RV free wall. 24-hour Holter monitoring recorded a PVC burden of 719 beats, without episodes of non-sustained or sustained ventricular tachycardia.

Finally, the diagnosis of ACM was established according to the 2010 task force criteria (16). Within the next years, cardiomyopathy progressed, with a further decline in LV function (LVEF 40 %) and an increase in RV size. On Holter-monitoring, PVC-burden increased to 2 %, and occasional non-sustained ventricular tachycardia was observed. Heart failure therapy was initiated (at the last follow-up, the patient was receiving guideline-directed medical therapy including an ACE inhibitor, eplerenone, dapagliflozin, and metoprolol) and an ICD was implanted as a primary prophylactic measure. Panel testing revealed homozygosity for the c.523+2T>C variant in *DSG2*. LVEF improved to 50 % with medical therapy. Clinically, the patient experienced a slight reduction in exercise capacity but maintained an overall good quality of life and stable clinical progression during follow-up.

However, approximately one decade after the initial diagnosis, the patient suddenly developed left hemiplegia, accompanied by weakness in the left arm and face. Cranial MRI revealed a M1 occlusion at the right site with a consequent infarct in the insular and lateral basal ganglia, corresponding to the clinical symptoms. The patient underwent intravenous thrombolysis and endovascular intervention with complete reperfusion without complications. TOE confirmed the known RV dilatation with reduced function and slightly reduced LV systolic function. A small LV-apical aneurysm was observed, but no thrombus was present (**Figure 1**, **Supp. Video 3A, B).** No patent foramen ovale was detected, and CT of the heart showed no thrombus.

Despite the absence of intracardiac thrombus on the CT scan (which was performed after systemic lysis), therapeutic anticoagulation was recommended, as the suspected embolic source—based on experience from the first two described cases—was a thrombus dissolved by lysis in the area of the apical aneurysm.

### Haplotype analysis revealed *DSG2* c.523+2T>C variant as founder mutation

To evaluate if the *DSG2* variant identified in all three patients is derived from a common founder, haplotype analysis was performed. Here, two regions of homozygosity (ROH) of 4.91 Mb total length on chromosomes 4 and 18 were identified common to all three patients (**Figure 2**). It must be noted that for patient 3 clinical exome sequencing was performed (and not a whole exome sequencing (WES) as for patients 1 and 2), which contains approximately one fifth of the number of genes of a WES. Therefore, the number of identical ROH shared by all three patients is most likely underestimated. The ROH on chromosome 18 is encompassing the *DSG2*-gene (**Figure 2A**), indicating an identical haplotype in all three patients. This suggests that the described *DSG2*-variant is a founder variant.

**Figure 2.**
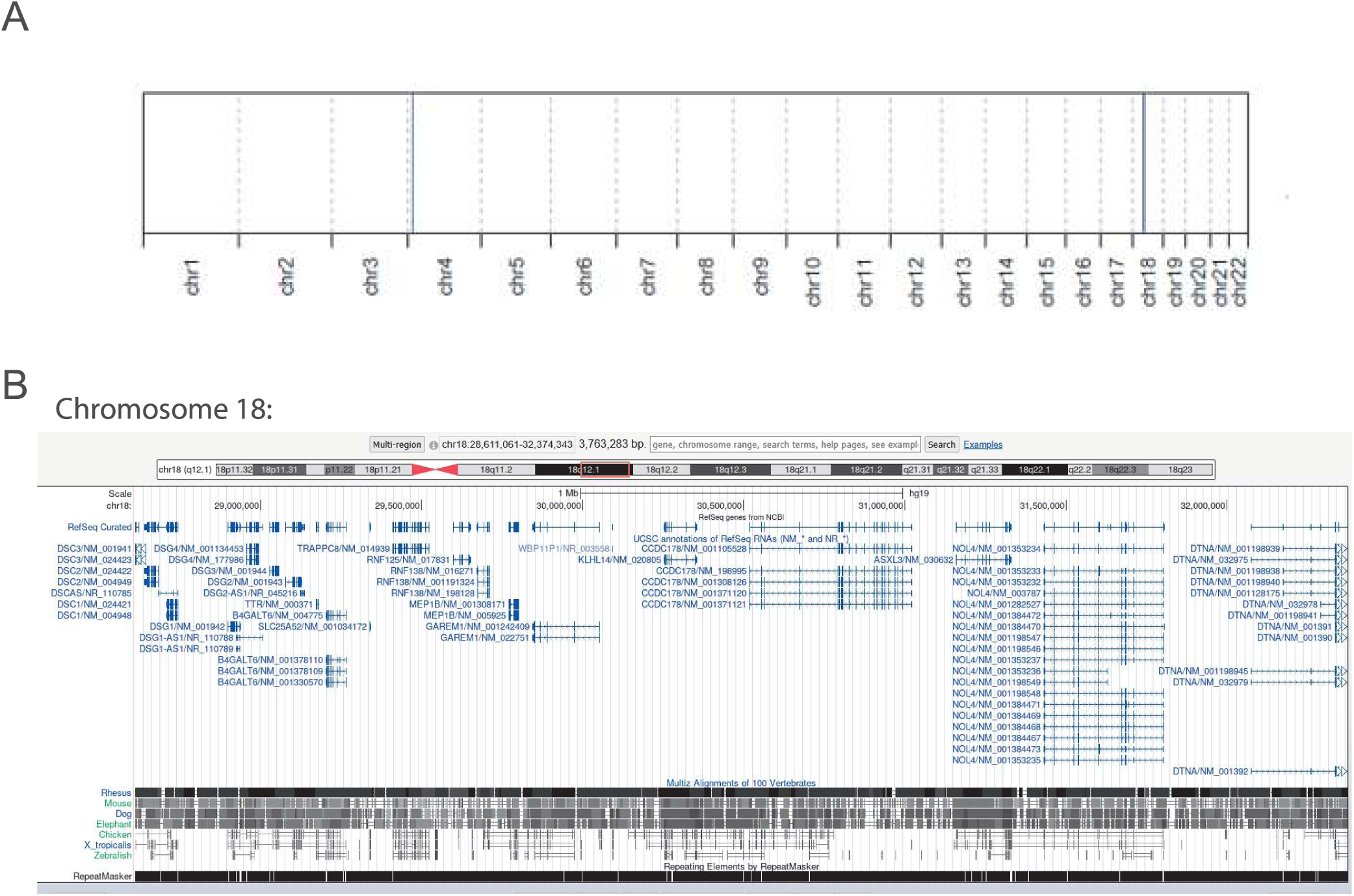
Haplotype analysis in patients 1-3. (A) Output of the AutoMap analysis. For patient 1 and 3 whole exome sequencing (WES) data and for patient 3 clinical exome sequencing data were used. Blue lines indicate a shared identical region of homozygosity (ROH), with the blue line on chromosome 18 encompassing the *DSG2*-gene. (B) Zoomed-in region of the genomic homozygous region of the *DSG2*-gene.

All three homozygous patients originate from a certain small geographic region in Switzerland with approximately 14,000 inhabitants. Thus, assuming a Hardy-Weinberg equilibrium (21), this would correspond to an estimated heterozygous carrier frequency of approximately 1 in 34 individuals. The *a priori* risk of an affected offspring from two individuals from this region is therefore 1:4,600. This estimate is likely conservative, given the probable under ascertainment of affected individuals in the region, as not all patients are likely diagnosed and, even when identified, may not all be followed at our center.

### Characterization of *Dsg2* mouse models to evaluate the patients’ phenotype

To analyse this newly described ACM phenotype more in detail, we evaluated two different Dsg2 mutant mouse models (**Figure 3A-C**): (1.) The Dsg2Δex5 model employs the *loxP*/Cre-system by crossing a previously described Dsg2-flox model, which bears a *Dsg2* gene with exon 5 being flanked by *loxP* sites (floxed) (18, 19), with the established CAG-CreER^T2^-strain expressing the Cre recombinase transgene under a ubiquitous tamoxifen-inducible promotor (004682, The Jackson Laboratory, Bar Harbor, USA). After administration of tamoxifen to the adult *Dsg2*^flox/flox^;Cre^Tg^ mice for 5 days, Cre expression is induced and exon 5 of *Dsg2* is deleted by the recombinase. The exon deletion leads to a frameshift and preterminal stop-codon with loss-of-function and depletion of the Dsg2 protein (**Figure 3D**). Thus, the Dsg2Δex5 model aims to mimic the described patient cases as the recombination induces an aberration in the similar genetic region resulting in a frameshift and truncation downstream of exon 5. *Dsg2*^flox/flox^ animals treated with tamoxifen without Cre transgene served as control group. (2.) To compare aberrations in the region of exon 5, which encodes parts of Dsg2 extracellular domain 2, with a distinct functional region of the protein, we employed the previously described Dsg2-W2A knock-in model (15) (**Figure 3B, C**). This point mutation at the binding interface of Dsg2 renders the protein adhesion-deficient leading to dysfunction of the desmosome while preserving the proteins’ membrane localization. Thus, this model enables the assessment of the consequences of loss of the adhesive function of Dsg2 without depletion of the protein (**Figure 3D, Supp. Figure 1A**).

**Figure 3.**
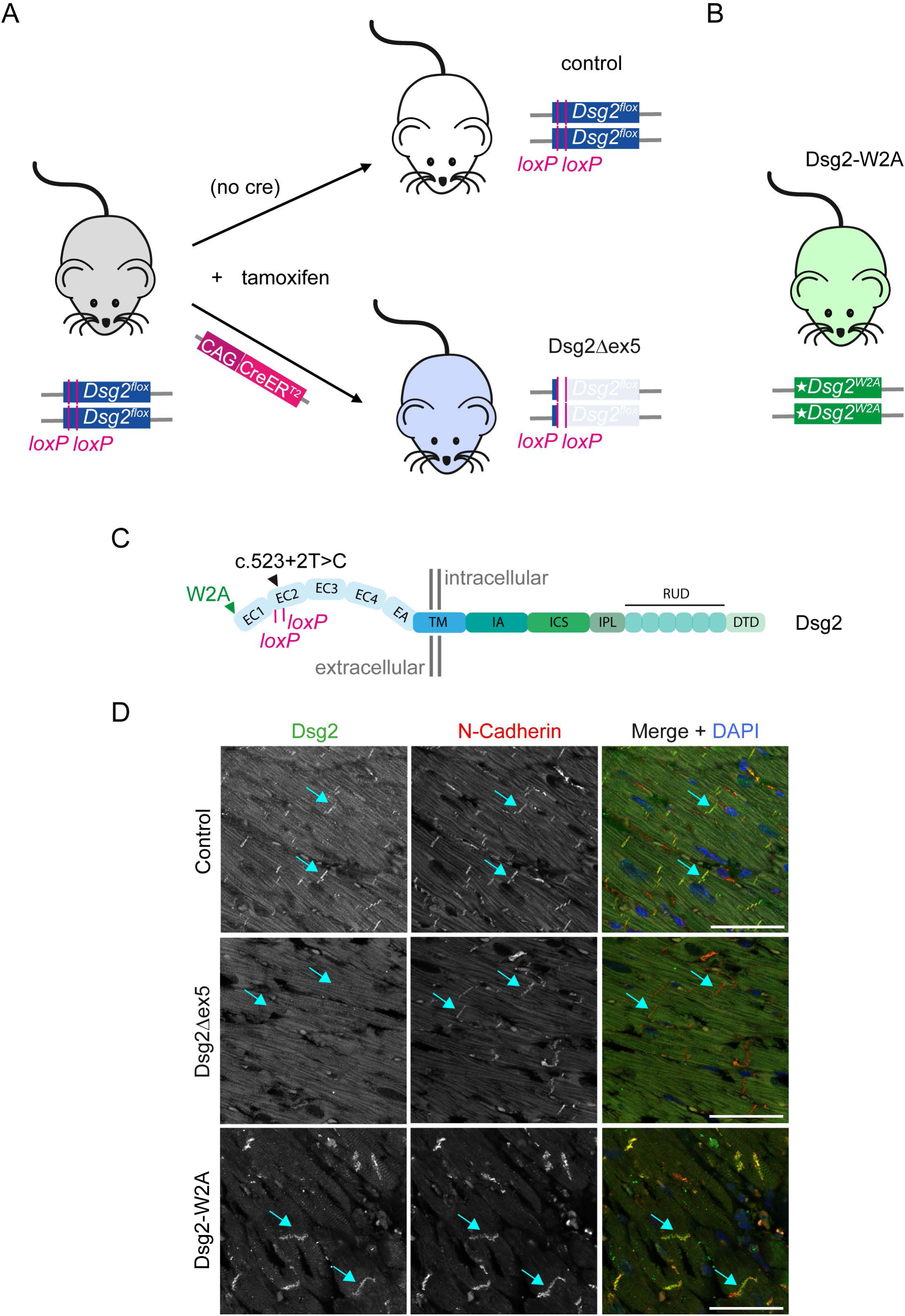
Overview of employed *Dsg2* mouse models. (A) To mimic the patient variant, a tamoxifen-inducible Dsg2 mutant mouse model (Dsg2Δex5) was evaluated. Exon 5 of *Dsg2* is flanked by two *loxP* sites (pink). Expression and activation of CAG-CreER^T2^ transgene by tamoxifen administration leads to deletion of exon 5 and depletion of Dsg2 protein mimicking the patients’ situation. Cre-negative mice treated with tamoxifen served as controls. (B) The Dsg2Δex5 model was compared to the adhesion-deficient Dsg2-W2A knock-in mutation described and established earlier in (15). (C) The schematic indicates the position of the *loxP* sites, the W2A mutation, and the location of the patient mutation with respect to the protein domains of Dsg2. The extracellular domains 1-4 (EC1-4), extracellular anchor (EA), transmembrane domain (TM), intracellular anchor domain (IA), intracellular catenin binding site (ICS), intracellular proline-rich linker (IPL), repeated unit domain (RUD), and DSG-terminal domain (DTD) are indicated. (D) Paraffin sections of murine hearts were stained for Dsg2 and N-Cadherin as intercalated disc marker (arrows indicate exemplary intercalated discs) and analysed by confocal imaging. DAPI was used as nuclei counterstain. Incubation with corresponding isotype IgG served as negative control and is shown in Supp. Figure 1A. Scale bar = 50 µm.

As a first step, the cardiac phenotype in these mouse models was evaluated in analogy to the clinical evaluation of ACM patients including echocardiography and ECG analysis. In echocardiography measurement 8 - 11 weeks post-induction, Dsg2Δex5 animals presented with reduced ventricular output (**Figure 4A**, control versus Dsg2Δex5 **Supp. Videos 4A,B** and **Supp. Videos 5A, B,** respectively) mainly affecting the RV (detected as lower ejection fraction, RVEF) (**Figure 4B**). This was accompanied by a mildly reduced LV ejection fraction. In two out of three Dsg2Δex5 animals, localized left ventricular apical wall thinning and movement irregularities were detectable indicating an LV aneurism (**Supp. Videos 5A, B**). In comparison, Dsg2-W2A animals at the stage of advanced disease (age of 6-12 months) exhibited impaired output function of both ventricles with reduced LVEF, RVEF, and TAPSE (**Figure 4, Supp. Videos 6A, B**). ECG analysis with recordings for 5 min revealed low voltage QRS complex morphology with reduced RS amplitude mainly driven by reduction of the R amplitude in Dsg2Δex5 mice and reduction of the S amplitude in Dsg2-W2A animals, while the QRS interval and other main parameters were not significantly altered (**Figure 5A, B, Supp. Figure 2**). Moreover, ventricular arrhythmias were detected in both models with all Dsg2Δex5 and half of the Dsg2-W2A animals presenting with premature ventricular contractions (PVCs) and partially non-sustained ventricular tachycardia (**Figure 5C - E**).

**Figure 4.**
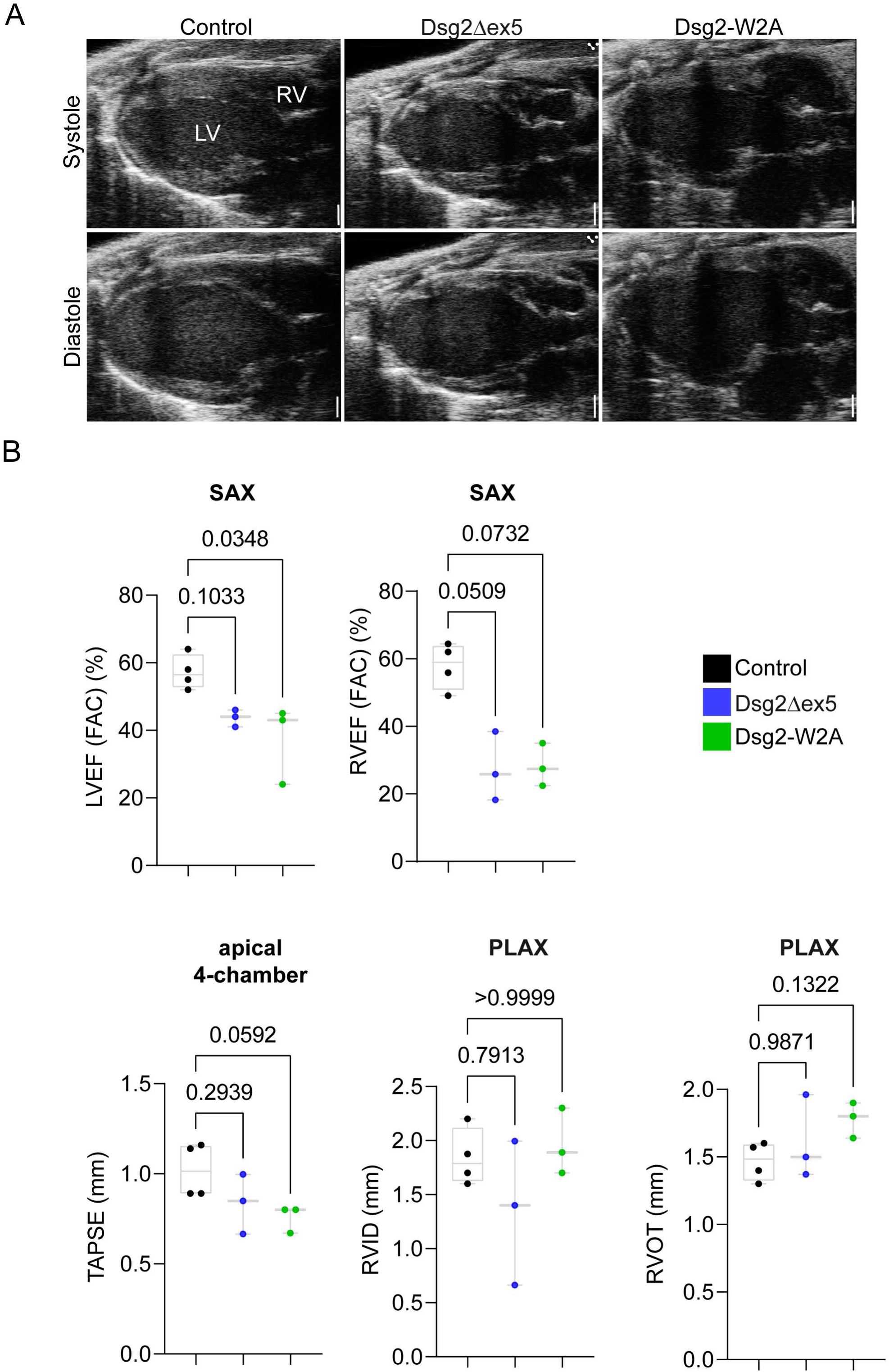
Impaired cardiac output function in Dsg2Δex5 and Dsg2-W2A mice with varying ventricular dominance. (A) Representative images of echocardiography measurements of control, Dsg2Δex5 and Dsg2-W2A hearts in PLAX view with indication of the right and left ventricle (RV, LV). Scale bar = 1 mm. (B) Analysis of the left and right ventricular ejection fraction (LVEF, RVEF) as fractional area change (FAC) in PLAX view, Tricuspid Annular Plane Systolic Excursion (TAPSE) in 4-chamber view, right ventricular outflow tract (RVOT) in PLAX and RV inner diameter (RVID) in diastole in PLAX view. Each data point represents one animal, p-values are indicated.

**Figure 5.**
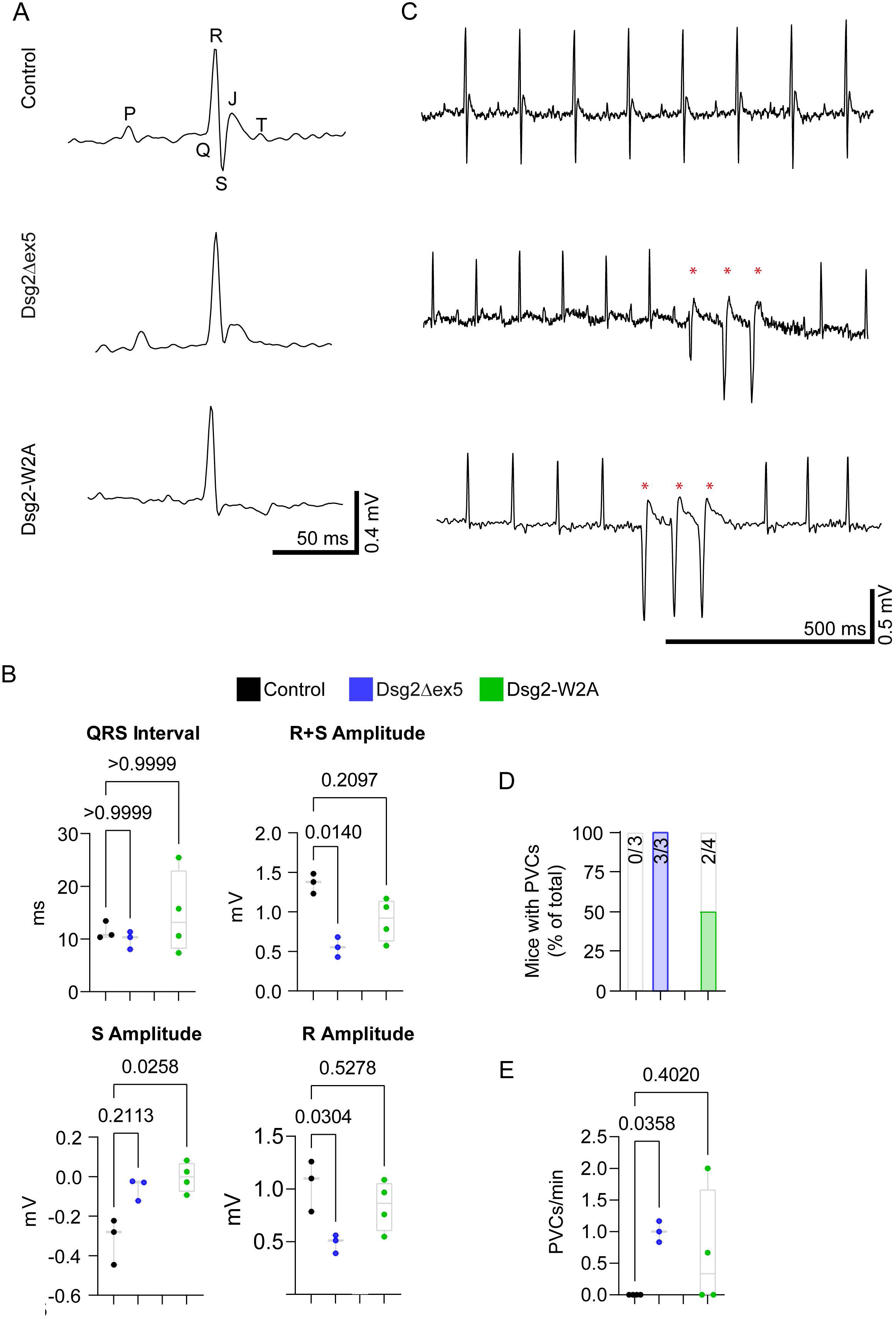
Altered electrical conduction and ventricular arrhythmia Dsg2Δex5 and Dsg2-W2A mice. (A) Representative electrocardiogram (ECG) traces showing QRS complex morphology with respective peaks indicated with (B) corresponding analysis of QRS interval, R+S amplitude, S amplitude, and R amplitude. For additional ECG parameters see Supp. Figure 2. (C) Representative ECG recording traces. Asterisks indicate premature ventricular contractions (PVCs). (D) Number of mice presenting with episodes of PVCs during a recording time of 5 minutes. Absolute animal numbers are indicated. One Dsg2-W2A mouse presented additionally with atrial fibrillation (not shown). (E) Quantification of PVCs per minute and animal. Each data point represents one animal, with p-values and scale bars indicated.

Following the functional analysis, hearts were dissected and collected for detailed histological work-up. Here, Dsg2Δex5 hearts demonstrated macroscopically visible RV dilatation and fibrotic patches, while Dsg2-W2A hearts exhibited a more pronounced phenotype with partial tissue calcifications (**Figure 6A**). Sirius red fibrosis staining of the hearts at different transversal levels (apex, apical, mid-papillary) revealed in Dsg2Δex5 mice a fibrotic pattern with collagen patches wrapping around the heart from the apex into the direction of the basis following the orientation of the cardiac muscle fibers. Large fibrotic areas were detected at the apical and the mid-papillary level, especially in the free wall of the RV and the interventricular septum (**Figure 6B - D**). The Dsg2-W2A animals exhibited a slightly distinct fibrotic phenotype, with pronounced fibrosis of the LV and RV free walls with only partial affection of the apical region and myocardial calcifications as a sign of very advanced disease. In summary, Dsg2Δex5 mice presented with an arrhythmogenic phenotype with RV-dominant functional impairment and biventricular fibrosis affecting the apex of the heart, similarly as observed in human patients, while the Dsg2-W2A model is partially deviating with minor apical fibrosis and pronounced biventricular functional impairment.

**Figure 6.**
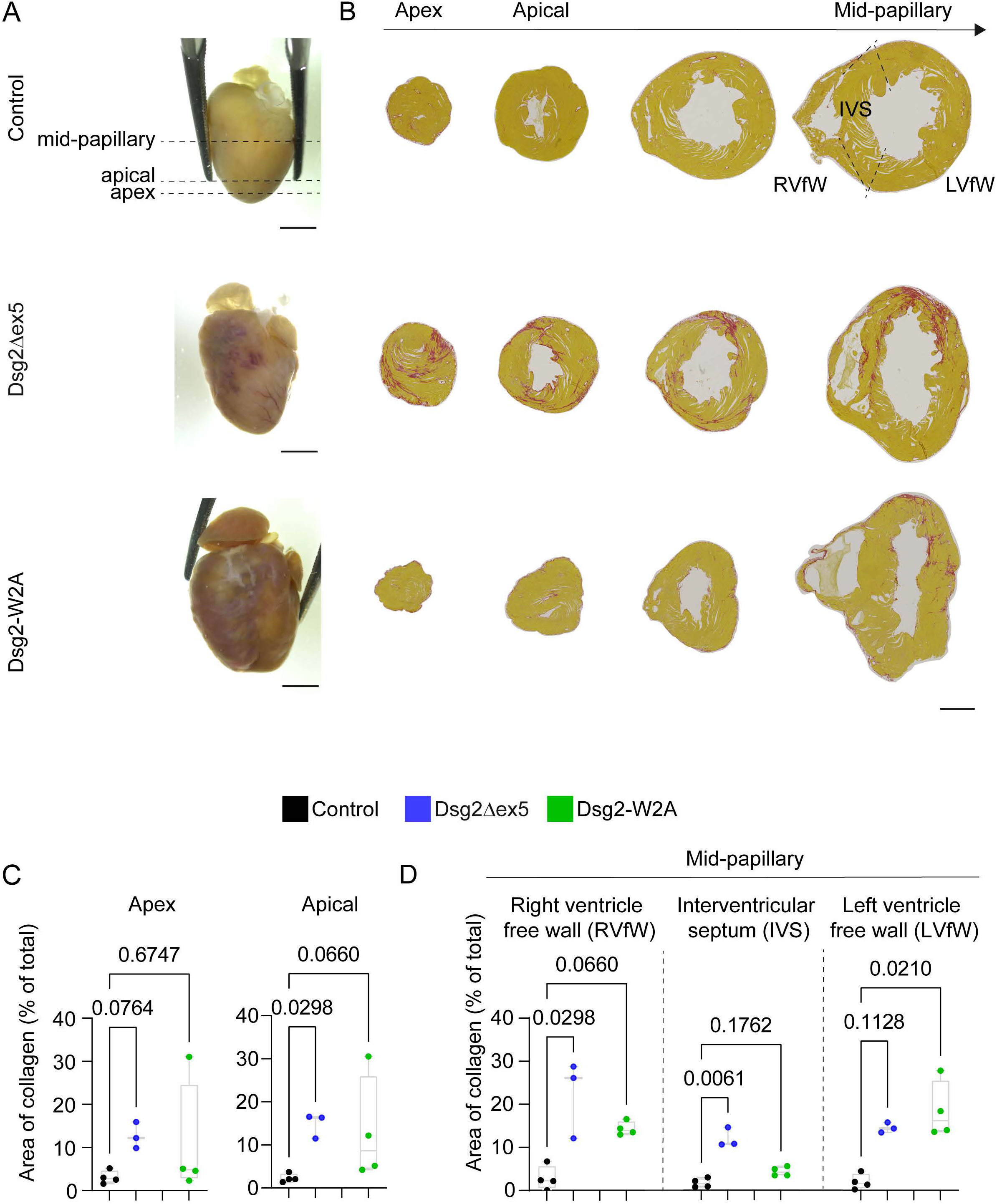
Biventricular fibrosis with an apical and right ventricular focus in Dsg2Δex5 mice versus a left ventricular focus in Dsg2-W2A animals. (A) Representative images of gross morphology of dissected hearts showing fibrosis and ventricular wall thinning. Scale bar = 3 mm. Histological assessment of collagen deposition by Sirius Red staining with representative images of paraffin sections in (B) and corresponding analysis in (C) and (D). The heart was cut in transversal sections and the fibrotic area was evaluated at the apex, the apical region and the midpapillary level with annotation and separate analysis of the right ventricular free wall (RVfW), the interventricular septum (IVS) and the left ventricular free wall (LVfW) as indicated, scale bar = 1 mm. Each data point represents one animal, p-values are indicated.

### Detection of LV thrombi in the Dsg2Δex5 mouse model

To evaluate the murine hearts for the presence of thrombi as detected in the patients, histological sections of the respective animals were stained for hematoxylin/eosin and assessed with respect to ventricular blood clots. Here, large RV clots were detectable in all three Dsg2Δex5 hearts with one animal showing also small LV clots. In one of the analysed Dsg2-W2A hearts, a large RV clot was visible. None of the control animals showed a blood clot (**Figure 7**). To determine if the identified structures are thrombi, which were present already in vivo and not post-mortem developed blood clots, the sections were stained with the thrombus marker fibrin, which confirmed large RV thrombi in all Dsg2Δex5 mice and the Dsg2-W2A heart with an additional small LV thrombi in one Dsg2Δex5 animal (**Figure 7, Supp. Figure 1B**). Moreover, during the evaluation period of 11 weeks post-induction, two Dsg2Δex5 animals were found dead in their cages. The autopsy revealed for one animal brain tissue disintegration within the left hemisphere potentially indicating an ischemic stroke (**Supp. Figure 3**).

**Figure 7.**
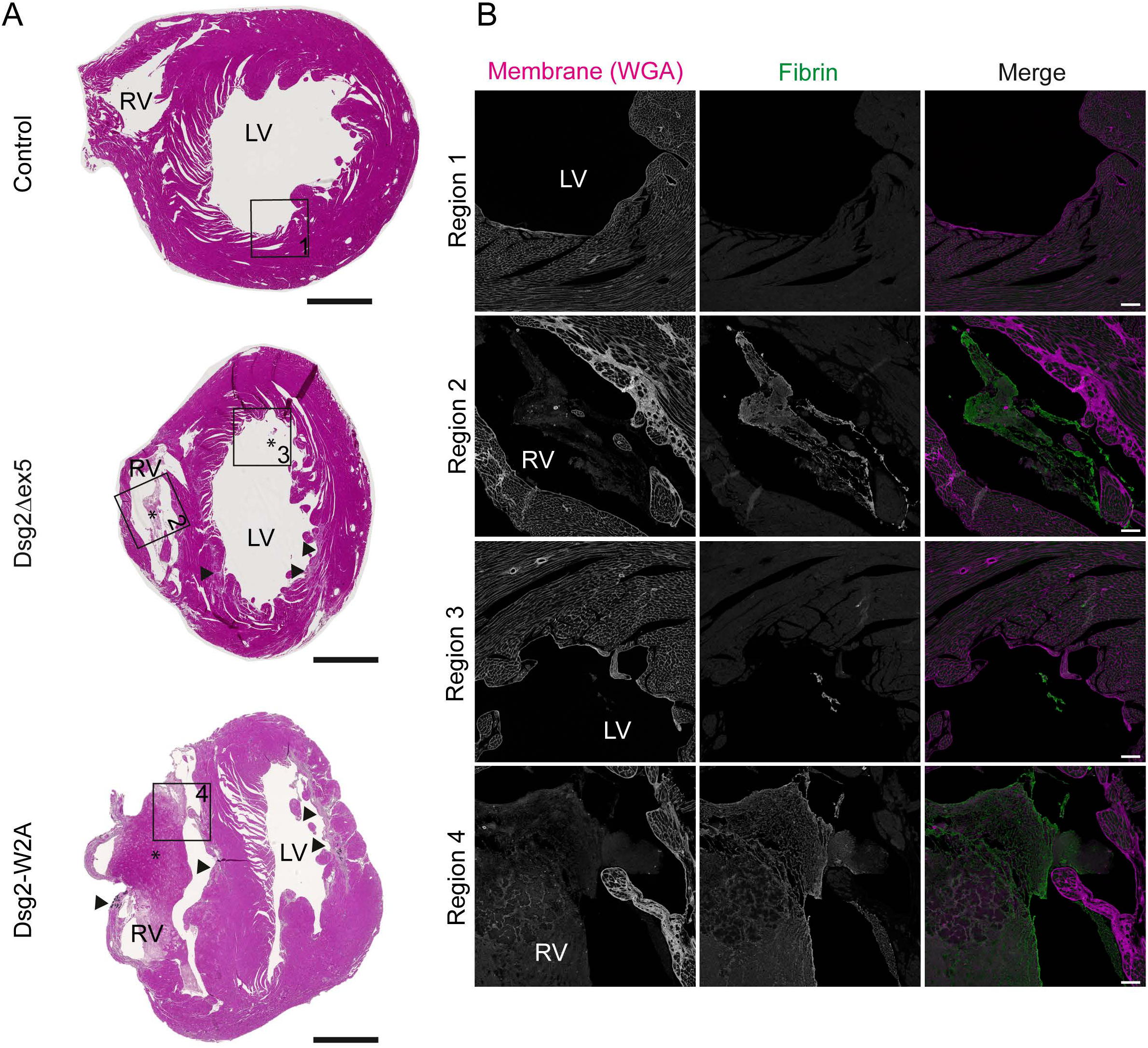
Occurrence of ventricular thrombi in Dsg2Δex5 and Dsg2-W2A hearts. (A) Overview images of heart paraffin sections stained for H.E.. Asterisks label ventricular thrombi, arrowheads point to fibrotic regions. Scale bar = 1 mm. (B) Adjacent paraffin sections were stained for the thrombus marker fibrin and analysed by confocal imaging. Localization of respective regions is indicated by rectangles in (A). Wheat-Germ-Agglutinin (WGA) staining of the cell membrane was used as a counterstain to mark cardiac tissue. Right ventricle (RV), left ventricle (LV). Incubation with corresponding isotype IgG served as negative control and is shown in Supp. Figure 1. Scale bar = 100 µm.

### Dsg2Δex5 mouse model resembles the phenotype of the Swiss founder mutation

In summary, the Dsg2Δex5 and Dsg2-W2A animals resemble central features of the three described patient cases. Particularly the Dsg2Δex5 mice presented with the same pathological features as the described patients including a low voltage ECG with occurrence of ventricular arrhythmia, dysfunctional cardiac output with a dominance for the RV, and fibrosis of the RV and LV apex accompanied with formation of an aneurism and (RV and) LV thrombus in some cases (**Table 1**). In contrast, the Dsg2-W2A mice were only partially resembling the patients’ features as the LV was functionally and structurally more severely affected, but without observation of a LV thrombus.

**Table 1.** Summary and comparison of data from the evaluated patients and mouse models. Not detected (n.d.), Right ventricular free wall (RVfW), interventricular septum (IVS), left ventricular free wall (LVfW). Check sign: present in all respective cases.

| Parameter and detection method |  | patient 1<br>♀ | patient 2<br>♂ | patient 3<br>♂ | Dsg2Δex5<br>model | Dsg2-W2A<br>model |
| --- | --- | --- | --- | --- | --- | --- |
| ECG | low voltage | ✓ | ✓ | ✓ | ✓ | reduced S peak |
| ECG | Ventricular arrhythmia | ✓ | ✓ | ✓ | ✓ | 2 out of 4 mice |
| TTE/MRI | Impaired RV function | ✓ | ✓ | ✓ | ✓ | (✓) |
| TTE/MRI | Impaired LV function | normal | Mild reduction | Mild reduction | Mild reduction | ✓ |
| TTE/MRI | LV apical aneurysm | ✓ | ✓ | ✓ | 2 out of 3 mice | n.d. |
| Imaging (MRI/SR) | LV apical fibrosis | ✓ | ✓ | ✓ | ✓ | 2 out of 4 mice |
| Imaging (MRI/SR) | Fibrosis in other regions | RVfW | RVfW | RVfW | RVfW, IVS | LVfW, (RVfW) |
| Imaging (MRI/SR) | Ventricular thrombus | LV: 5 x 5 mm | LV: 4 x 8 mm | None, after thrombolysis | RV: 3/3<br>LV: 1/3 | RV: 1/4<br>LV: 0/4 |
| Clinic/ post-mortem | Cardioembolic stroke | ✓ | n.d. | ✓ | potential stroke in one mouse | n.d. |

## Discussion

This study identifies LV thrombus formation as a novel pathological feature of DSG2-associated ACM. We report three unrelated patients homozygous for the Swiss *DSG2* founder variant c.523+2T>C who developed LV thrombi and/or presumed cardioembolic stroke. These thrombi were localized at an apical LV aneurysm caused by local fibrosis as detected by cardiac magnetic resonance imaging, despite preserved or only mildly reduced LV ejection fraction. These findings suggest that regional structural remodeling rather than global systolic impairment constitutes the critical substrate for thrombus formation.

To mechanistically evaluate the patient situation, we employed the Dsg2Δex5 mouse model, which bears an aberration in the same genetic region observed in carriers of the c.523+2T>C variant leading to depletion of the protein. In parallel, we studied the Dsg2-W2A knock-in model, in which desmosomal adhesion is selectively impaired through disruption of the Dsg2 binding mechanism with preserved membrane localization of the protein (15).

Both, Dsg2Δex5 and Dsg2-W2A mice, developed biventricular fibrosis with altered electrical conduction and ventricular arrythmia. In Dsg2Δex5, mainly the RV systolic output was reduced while Dsg2-W2A exhibited a global systolic dysfunction. However, pronounced apical involvement with fibrosis and aneurysm and left ventricular thrombi were specifically identified in Dsg2Δex5 hearts. These findings suggest that loss of *Dsg2* creates a particularly vulnerable structural substrate for intracavitary thrombogenesis.

In ECG analysis, *Dsg2*-mutant hearts demonstrated ventricular arrhythmias and reduced QRS amplitudes consistent with low-voltage ECG patterns similar to the patients. The concordance of low-voltage signals with extensive fibrotic remodeling reinforces the concept that myocardial replacement fibrosis in DSG2-associated ACM establishes both an arrhythmogenic and prothrombotic milieu. Apical-predominant fibrosis and regional wall motion abnormalities likely promote localized blood stasis, providing a mechanistic link between structural remodeling and thrombus formation independent of advanced global systolic dysfunction.

Taken together, our translational approach allows discrimination between the consequences of genetic aberrations in the region of exon 5 with complete DSG2 deficiency and selective dysfunction of the adhesive interface. The data indicate that homozygous *DSG2* loss-of-function drives a distinct fibrotic–thrombotic phenotype characterized by apical fibrosis, electrical instability, and LV thrombus formation. To our knowledge, this represents the first description of LV apical fibrosis associated with thrombus formation as a variant-specific manifestation of ACM.

To better understand these findings, it is important to consider the established role of *DSG2* in the pathogenesis of ACM, which was initially described in 2006 (22, 23). This genetic association was subsequently corroborated by functional studies in a mouse model published in 2009, which provided in vivo evidence of *DSG2*’s critical role in maintaining cardiac desmosomal integrity and its contribution to ACM pathophysiology (24). Since then, *DSG2* variants have been identified in approximately 4 – 10 % of ACM patients in western countries, with a higher prevalence of around 15 % reported in Asian cohorts (6–8, 10). Already in 2009, an accumulation of biallelic or digenic *DSG2* mutations was observed in cohort studies, suggesting early recognition that bigenic or recessive inheritance might play an important role in *DSG2* associated ACM (6). In line with this, a particular variant — *DSG2* p.(Phe531Cys) — was later identified as a regional founder mutation in China. This variant demonstrates full disease penetrance in the homozygous state, whereas heterozygous carriers are typically asymptomatic or present only with mild clinical features, further underscoring the relevance of population-specific genetic backgrounds in modulating disease expression (10, 12). Building on these earlier insights, a recently published cohort study has further highlighted the complexity of the genetic landscape in *DSG2/DSC2*-associated ACM. It is reported that a substantial proportion of affected individuals (28.8%) carry multiple pathogenic or likely pathogenic variants in desmosomal cadherin genes, particularly in the Chinese population. Patients with multiple variants were diagnosed with ACM at a younger age and experienced higher rates of end-stage heart failure and malignant ventricular arrhythmias compared to those with a single variant (11). Our findings of severe disease associated with homozygosity of a *DSG2*-variant despite completely unremarkable family histories align with these observations. A single case report has previously described the variant identified in our cohort, albeit in a different clinical context, namely inflammatory features in homozygous *DSG2* cardiomyopathy mimicking cardiac sarcoidosis (25). To our knowledge, the description of the Swiss founder variant represents the first report of a *DSG2* founder mutation outside of China - although recent cohort data also suggest the presence of homozygous *DSG2* patients within western populations (10, 11).

Emphasizing the influence of population-specific genetic backgrounds, it is crucial to understand how these genetic variations manifest in cardiac pathology. Fibrosis is a hallmark of ACM and is also included as a criterion in its diagnostic guidelines (3). Data from the largest *DSG2* cohort revealed LGE in approximately 72 % of patients. Among the cases with biventricular involvement, LV LGE predominantly affects the subepicardial layer of the lateral and posterior walls, occurring in 70.5 % of these patients (11). Our clinical and mouse model data, however, highlight a distinct phenotype associated with the Swiss *DSG2* founder variant, where fibrosis is predominantly localized to the LV apex. This apical fibrosis appears to represent a variant-specific phenotype, observed in homozygous but not heterozygous carriers, and differs from the more widespread fibrotic patterns commonly reported in DSG2-associated ACM populations.

In contrast to the frequently described and clinically well-characterized fibrosis, LV thrombus formation is uncommon in ACM. However, RV thrombi have been described as a rare but recognized complication in previous studies. Reported in approximately 2 – 4 % of ACM patients, RV thrombi typically occur in the context of severe RV dysfunction or advanced disease (13, 14). In a cohort of 193 ACM patients, thrombi were documented in 10 individuals (4.1 %), with nine located in the RV and a single LV apical thrombus identified in a patient with severely reduced LVEF — though no genetic data were available (14). Similarly, in a larger cohort of 467 ACM patients, 13 (2.8 %) developed RV thrombi, and four of these also had LV thrombi; however, all four had significant comorbidities, including ischemic cardiomyopathy, hypertrophic cardiomyopathy (HCM), dilated cardiomyopathy (DCM), or the presence of a left ventricular assist device (LVAD) (13). To our knowledge, the aforementioned patient with severely impaired LVEF in the first cohort remains the only published case of isolated LV thrombus formation in ACM in the absence of overt comorbidity or device therapy (14), while no cases have been described with thrombus formation despite normal or only mildly impaired LVEF. Against this background, the observation of three unrelated patients homozygous for the same *DSG2* founder variant — each presenting with left ventricular (LV) thrombi precisely localized to the apex and/or cardioembolic stroke despite preserved or only mildly reduced LVEF — is particularly striking. In all cases and consistently reproduced in the corresponding *DSG2* mouse model, myocardial fibrosis was confined to the LV apex, indicating a strong anatomical and pathophysiological association between focal myocardial injury and thrombus formation. Our results indicate that the identified *DSG2* founder variant exerts a variant-specific effect, primarily inducing apical fibrosis in the LV, thereby playing a unique pathogenic role in thrombus formation. The phenotypic consistency and unique localization of fibrosis and thrombi in our cases suggest that the homozygous *DSG2* founder variant may define a distinct cardiomyopathic and thrombotic subtype.

## Conclusion

In conclusion, our combined clinical and experimental findings define a novel and previously unrecognized phenotype of *DSG2*-related ACM characterized by biventricular disease with a predilection for LV apical fibrosis that predisposes homozygous carriers of the Swiss founder variant *DSG2* c.523+2T>C to left ventricular thrombus formation and cardioembolic stroke. Our translational data identify deficiency of DSG2 as the central pathomechanistic driver of this fibrotic–thrombotic phenotype, while partial phenotypic differences can occur for different DSG2 loss-of-function mutations. These observations highlight the necessity of precision medicine approaches in ACM that consider not only the affected gene but also the specific pathogenic mechanism and variant zygosity. Importantly, variants resulting in complete or near-complete loss of DSG2 protein function — independent of their precise genomic location — may confer a comparable risk for apical fibrosis and intracavitary thrombosis. Thus, the thromboembolic propensity observed here may extend beyond the Swiss founder variant to other *DSG2* loss-of-function mutations.

Accordingly, we have implemented a variant-informed management strategy in homozygous carriers, initiating prophylactic anticoagulation upon detection of LV apical fibrosis by cardiac MRI to mitigate thrombus formation and stroke risk. Collectively, these findings expand the clinical spectrum of *DSG2*-associated ACM, identify a previously underrecognized thromboembolic complication, and challenge current paradigms of risk stratification that are primarily centered on arrhythmic risk rather than structural–thrombotic vulnerability.

## Data Availability

All data produced in the present study are available upon reasonable request to the authors.

## Sources of funding

This work was supported by a Grant from the Gottfried & Julia Bangerter-Rhyner-Stiftung and a Grant from the Inselspital Bern (“Nachwuchsförderungs-Grant”) to MR. CS is funded by the Swiss National Science Foundation (#218454 Starting Grant); the Talent4Bern program of the Medical Faculty, University of Bern and the Heike und Wolfgang Mühlbauer Stiftung, Hamburg.

## Acknowledgements

The authors gratefully acknowledge the patients who provided consent for the publication of their clinical cases. We thank Arnd Heuser (Animal Phenotyping Platform, Max-Delbrück Center, Berlin) for providing the *Dsg2*-flox mouse line; Aude Zimmermann, Lea Galvagno, Anja Huzinker, Florian Sprich (all DBM, University Basel), Julia Kemmling and Jennifer Schröder-Schwarz (UKE Hamburg) for technical assistance; Alain Brühlhart and the team from the Animal Facility, DBM, University of Basel for animal care taking; and the Microscopy Imaging Facility (DFG Research Infrastructure Portal: RI_00489) at the University Medical Center Hamburg for technical microscopy support.

## Disclosures

The authors state that there is no conflict of interests.

## Abbreviations

ACM: Arrhythmogenic Cardiomyopathy
ARVC: Arrhythmogenic Right Ventricular Cardiomyopathy
CT: Computed Tomography
DSC2: Desmocollin-2
DSG2: Desmoglein-2
DSP: Desmoplakin
ECG: Electrocardiogram
EDV: End-Diastolic Volume
FAC: Fractional Area Change
ICD: Implantable Cardioverter Defibrillator
JUP: Junctional Plakoglobin (Plakoglobin)
LGE: Late gadolinium enhancement
LVEF: Left Ventricular Ejection Fraction
LV: Left Ventricle / Left Ventricular
MRI: Magnetic Resonance Imaging
PLAX: Parasternal Long Axis View
PKP2: Plakophilin-2
PVC: Premature Ventricular Contraction
ROH: Regions of Homozygosity
RV: Right Ventricle / Right Ventricular
RVOT: Right Ventricular Outflow Tract
SAX: Short-Axis View
TAPSE: Tricuspid Annular Plane Systolic Excursion
TOE: Transesophageal Echocardiography
TTE: Transthoracic Echocardiography
TV annulus DTI S’: Trikuspid valve annulus doppler tissue imaging systolic velocity
WES: Whole Exome Sequencing

---

**Supplementary Figure 1. Isotype control images corresponding to Figure 3 and Figure 7**. Confocal images of corresponding regions of interest are shown. Sections were incubated with isotype IgG to exclude positive signal due to unspecific binding of the secondary antibody. (A) Isotype control panel for Figure 3. scale bar = 50 µm. (B) Isotype control panel for Figure 9. WGA was applied as a membrane marker to counterstain cardiac tissue, DAPI was applied as nuclear counterstain. Right ventricle (RV), left ventricle (LV); scale bar = 100 µm.

**Supplementary Figure 2. Additional ECG parameters corresponding to Figure 5**. Analysis of heart rate, PR Interval, QT Interval, P Amplitude and Q Amplitude in addition to parameters quantified in Figure 5. Each data point represents one animal, p-values are indicated.

**Supplementary Figure 3. Single case of potential brain ischemia in Dsg2Δex5.** (A) Gross morphology of the unfixed brain of a Dsg2Δex5 mouse found dead. The box indicates damaged brain tissue in the left hemisphere. (B) Cryo-preserved cross section of the same brain for confirmation. Box indicates damaged brain tissue. Scale bars = 5 mm

**Supplementary Video 1.** Transthoracic echocardiographic (TTE) imaging in apical four-chamber view for patient 1.

**Supplementary Video 2.** (A, B) Transthoracic echocardiographic (TTE) imaging in two different angles in four chamber view of patient 2.

**Supplementary Video 3. Echocardiography imaging of patient 3** (A) Transesophageal echocardiography (TOE) imaging in the four chamber view for patient 3. (B) Transthoracic echocardiography (TTE) imaging in the parasternal long axis view for patient 3 with a focus on the apex.

**Supplementary Video 4. Transthoracic echocardiographic (TTE) of a control mouse.** (A) in the short axis view (SAX) and (B) the long axis (PLAX) view.

**Supplementary Video 5. Transthoracic echocardiographic (TTE) of a Dsg2Δex5 mouse.** (A) in the short axis view (SAX) and (B) the long axis (PLAX) view.

**Supplementary Video 6. Transthoracic echocardiographic (TTE) of a Dsg2-W2A mouse.** (A) in the short axis view (SAX) and (B) the long axis (PLAX) view.

## Notes

### Competing Interest Statement

The authors have declared no competing interest.

### Author Declarations

Cantonal Ethics Board Bern, Bern, Switzerland, 2021-01396 gave ethical approval for this work.

