## Supplementary figures and images for "A Swiss *DSG2* Founder Variant Promotes Left Ventricular Thrombus Formation Causing Cardioembolic Stroke in Autosomal Recessive Arrhythmogenic Cardiomyopathy"

### Supplementary Figure 1

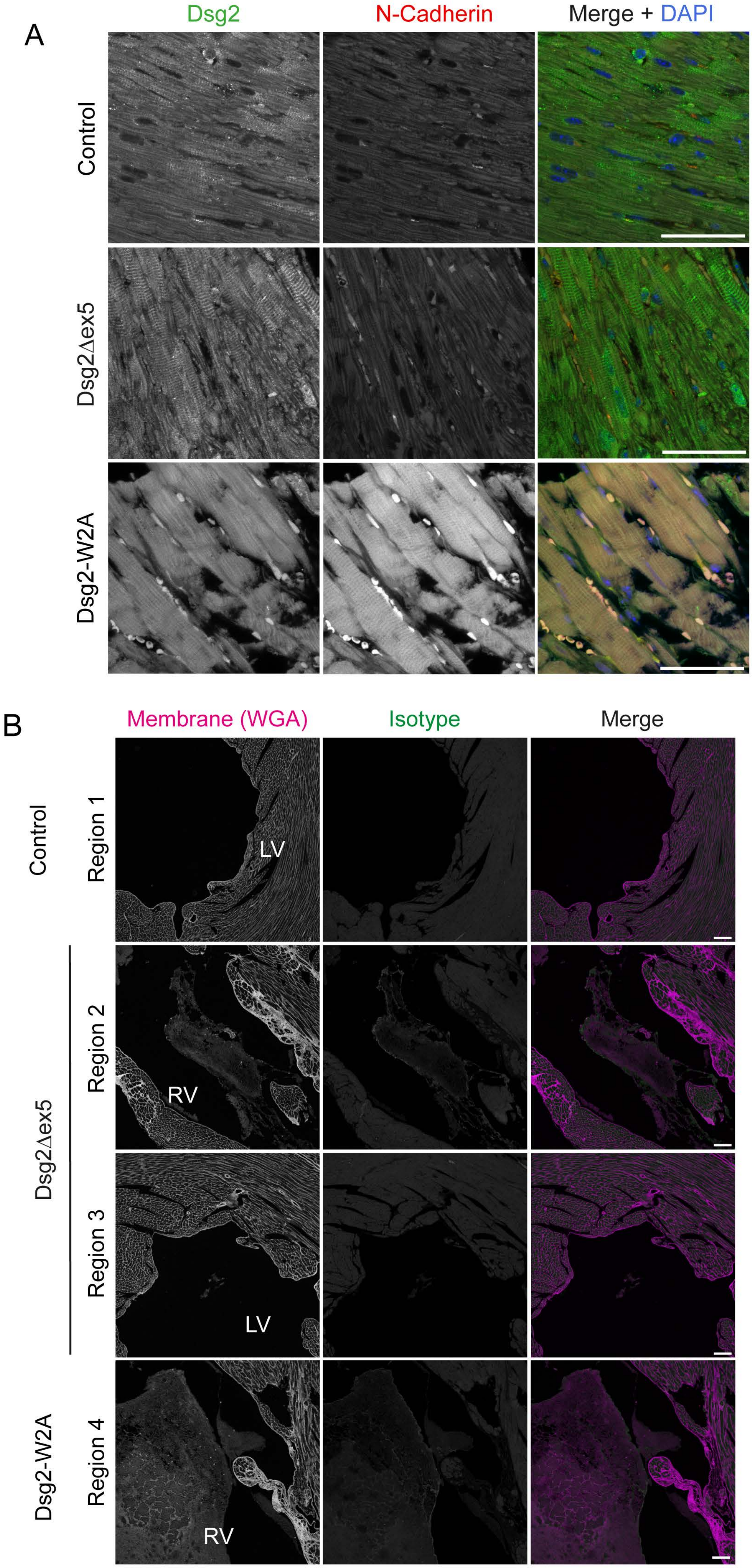


Supp. Figure 1

### Supplementary Figure 2

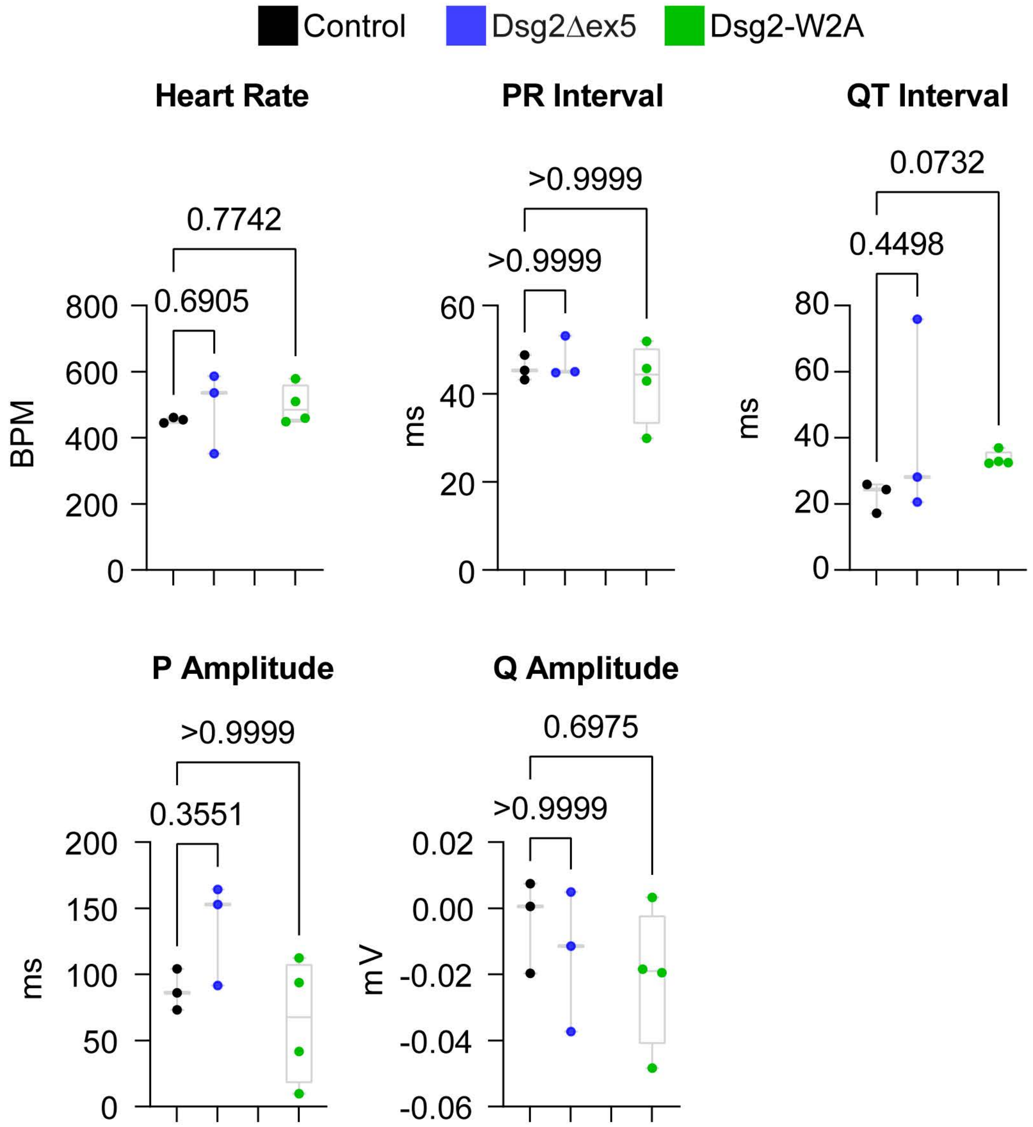


Supp. Figure 2

### Supplementary Figure 3

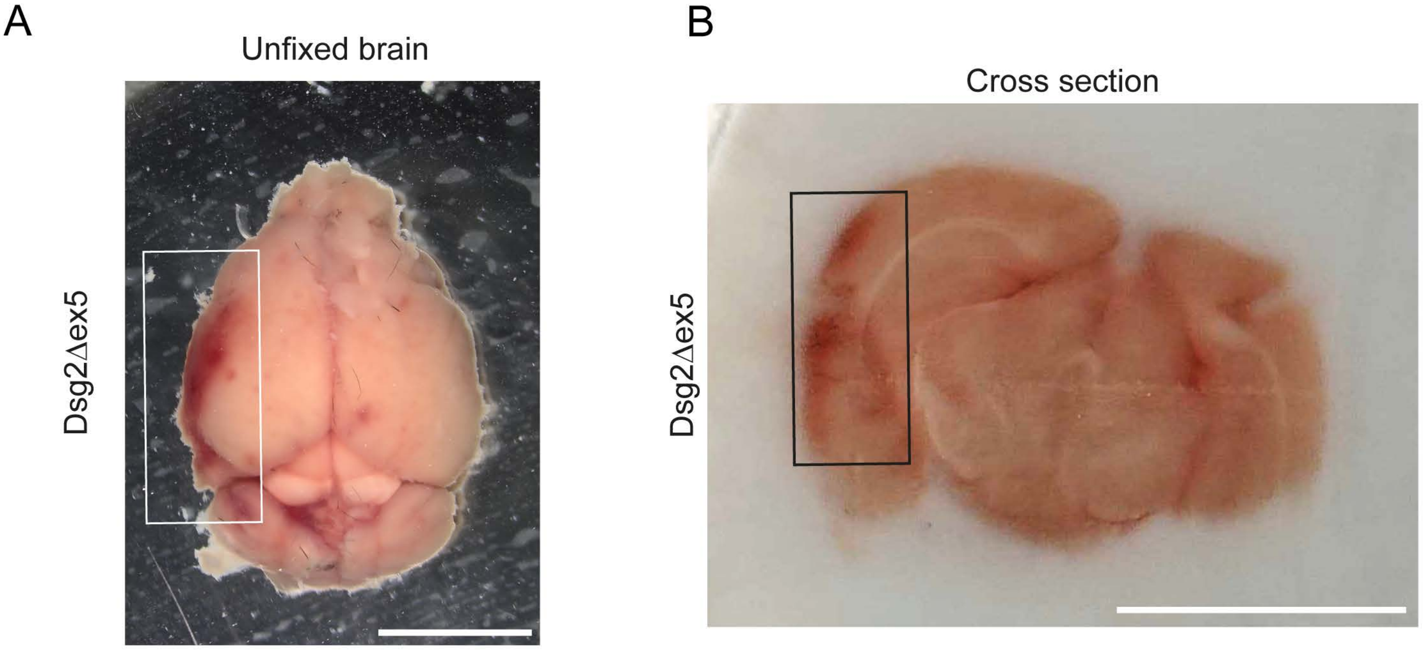


Supp. Figure 3
